# Altered sensorimotor-association axis patterning of global functional connectivity in an autism subtype with low levels of language, intellectual, and adaptive functioning

**DOI:** 10.1101/2025.10.30.25339153

**Authors:** Ines Severino, Veronica Mandelli, Natasha Bertelsen, Michael V. Lombardo

## Abstract

In early development autism can be stratified into subtypes differentiated by non-core language, intellectual, motor, and adaptive functioning features. In toddlerhood, these subtypes show different genomic patterning effects on brain structure and function that follow early primary axes of neurodevelopmental organization. This leads to the hypothesis that early cortical patterning differences may continue to be evident between subtypes in later development within hierarchical organization along the sensorimotor-association (S-A) axis. To test this hypothesis, we first demonstrate that unsupervised data-driven techniques can detect 2 autism subtypes (high versus low) with very high accuracy (92%) in late childhood to adulthood based on language, intellectual, and adaptive functioning features. In resting state fMRI data, we find that global functional connectivity is differentially patterned in the subtypes along the S-A axis. Autism with lower levels of language, intellectual, and adaptive functioning have hypo-connected sensorimotor and hyper-connected association areas relative to a non-autistic comparison group. These findings suggest that cortical patterning of global functional connectivity along the S-A axis differentiates an autism subtype with low levels language, intellectual, and adaptive functioning skills.

---

The current definition of autism, centered on early developmental difficulties in social-communication and restricted, repetitive behaviors [1], encompasses a wide spectrum of clinical presentations and previously distinct subtype labels [2]. While the diagnostic change from DSM-IV to DSM-5 may have helped to maximize consensus and reduce discrepancies associated with subtype labeling, it also had the side effect of deprioritizing features such as early language and intellectual functioning. Such changes may have hindered rather than helped research make progress towards isolating distinct neurobiological explanations behind different autisms. For example, meta-analysis has shown a time-dependent trend of attenuating effect sizes in case-control studies over time[3]. Such an effect highlights the need for stratification to make progress towards understanding heterogeneous neurobiology in autism. Since the change from DSM-IV to DSM-5, evidence has emerged indicating that early language, intellectual, motor, and adaptive functioning (LIMA) skills are key features that can be used to stratify autism in ways that both clinically and neurobiologically relevant [4–9]. Therefore, stratification approaches that target LIMA features may show promise in future research.

Our prior work has showed that stratification by early non-core LIMA features results in a type I vs type II distinction and that this distinction is sensitive to differences in patterned neural response to speech measured with fMRI [10]. We also found atypical genomic cortical patterning effects emerge and differentially explain task-related fMRI response to speech and cortical morphometric features like surface area and cortical thickness [6, 7, 10] . These early genomic patterning effects vary along important neurodevelopmentally sensitive anterior-posterior (A-P) and dorsal-ventral (D-V) axes [7] that are key to helping cortical areas develop their own unique identities[11]. Post-mortem gene expression studies in autism also identify atypical attenuation of genomic cortical patterning along the A-P axis [12–14]. This work suggests that genomically-mediated cortical patterning effects along key neurodevelopmentally sensitive axes may be key to explaining why autism can be differentiated into phenotypically distinctive LIMA subtypes.

As neurotypical brain development progresses, cortical patterning continues to be refined and shaped beyond the A-P and D-V axes and into a hierarchical organization that can be identified across multiple modalities that distinguishes sensorimotor versus association areas (i.e. the sensorimotor-association (S-A) axis) [15–18]. Resting state fMRI (rsfMRI) functional connectivity differentiation along the S-A axis is reduced in autism, with difficulty for association networks such as the default mode network to act as a functional ‘sink’ of information flow from sensory areas [19]. Mosaic patterns of functional hypo-connectivity between sensory and hyper-connectivity between associative regions have also been observed in large-scale rsfMRI studies [20, 21]. At a cognitive level, atypical patterning along the S-A axis in autism may be helpful in explaining imbalances between exteroceptive (sensory) and interoceptive (association) processing in autism [22]. Thus, examination of the S-A axis at later points in development may be important for identifying mechanisms that may differentially underpin autism LIMA subtypes.

In this study, we first tested whether distinctive LIMA subtypes can be identified in autism in the age range of mid-to-late childhood to adulthood. We first show that a phenotypic stratification model can be built and applied to neuroimaging datasets sampling primarily individuals older than 68 months. This stratification model uses unsupervised clustering on standardized clinical measures of language, intellectual, and adaptive functioning (LIAF) features taken from intelligence tests and adaptive functioning measures. The ‘M’ in the non-core ‘LIMA’ features for ‘motor’ functioning was not included in this work primarily due to the lack of sampling motor abilities by standardized intelligence and adaptive functioning tests at older ages that overlap with neuroimaging samples. Second, given the ability to identify such LIAF subtypes, we hypothesized that these subtypes will be differentiated in global functional connectivity along important neurodevelopmentally sensitive

cortical patterning axes such as the S-A axis. We used resting state fMRI data and computed a metric of global functional connectivity called weighted global brain connectivity (wGBC). wGBC is a regional summary measure indicative of the average functional coupling of a given region with the rest of the brain. In graph theory, this measure is equivalently known as weighted degree centrality. Past work on the unweighted version of this metric (i.e. unweighted degree centrality) has shown reproducible case-control differences in autism that manifest as both hypo- and hyper-connectivity [20]. Furthermore, rsfMRI work in animal models with known genomic causes of autism have identified numerous differences in wGBC (e.g., [23–25]) Thus, wGBC serves as a good baseline starting point for examining atypical neural connectivity at global levels in autism subtypes.

## Methods and Materials

### NDA dataset

The primary dataset used in this work for the purposes of building a generalizable stratification model for autistic individuals older than 68 months comes from the National Institute of Mental Health Data Archive (NDA) (https://nda.nih.gov/). NDA comprises multiple datasets containing standardized clinical measures of intellectual functioning, including the Wechsler Scales and the Differential Ability Scales (DAS), as well as adaptive functioning assessments using the Vineland Adaptive Behavior Scales (VABS) [26, 27]. Different versions of the Wechsler Scales were used depending on age group: the Wechsler Intelligence Scale for Children (WISC) [28–30] for individuals aged 6 to 16 years, the Wechsler Adult Intelligence Scale (WAIS) [31, 32] for those older than 16 years, and their abbreviated Wechsler Abbreviated Scale of Intelligence (WASI-II) [33]. NDA also includes two versions of the Differential Ability Scales–Second Edition (DAS-II) [34]: the Early Years form for children aged 2.6 to 6.11 years, and the School-Age form for children aged 7 to 17 years.

All relevant Wechsler, DAS, and VABS data were downloaded from NDA in July 2023. This included cognitive (WISC-III, IV, and V; WAIS-III and IV; and WASI-II; DAS-II Early Years and School-Age forms, n = 42,373) and VABS adaptive functioning assessments (Second Edition— Parent/Caregiver or Survey form, and Third Edition; n = 31,061). Only data from individuals aged above 68 months through adulthood were retained. From the VABS, standard scores for the communication, daily living skills, and socialization domains were extracted for further analysis. For intellectual ability, we extracted verbal and non-verbal (i.e., perceptual reasoning) quotients (see Supplementary Table 2 for detailed variable mappings). Next, we implemented a series of data cleaning steps that entailed selecting only individuals with an autism diagnosis, dropping duplicate data, extracting the earliest available time point above the age of 68 months, selecting only individuals whose IQ and VABS data were assessed not more than one year apart, and finally dropping individuals with more than one missing subscale for each of the instruments used. Finally, only standardized scores from VABS and IQ assessments were retained for clustering. After data cleaning and merging of IQ and VABS scores, a total of n=419 autistic individuals remained for further downstream analysis (mean age = 12.6 years ± 5.7, age range = 5.8–35.9, % female = 17.4%, mean FIQ = 98.23 ± 20.5) (see Supplementary Table 1 for NDA GUIDs and collection IDs of the participants included).

### Resting state fMRI datasets

To evaluate differences in global functional connectivity across autism subgroups, we utilized data from two independent large-scale consortia aggregating neuroimaging and phenotypic data from individuals with autism and neurotypical controls: (1) the Autism Brain Imaging Data Exchange (ABIDE) [35, 36] and (2) the Gender Exploration of Neurogenetics and Development to Advanced Autism Research (GENDAAR) initiative [37, 38]. Autistic participants were included if they had available measures of verbal and non-verbal IQ and adaptive functioning. From ABIDE I and II datasets, we specifically utilized data from the New York University site (NYU, NYU1, NYU2) as this was the only site that had all the requisite behavioral data to utilize in phenotypic stratification. After behavioral data cleaning and variable filtering there were a total of n=143 autistic individuals (mean age ± SD = 11.9 ± 6.9 years; age range = 5.1–39.1; 12.6% female; mean FIQ ± SD = 106 ± 17) and n=135 typically developing (TD) controls (mean age ± SD = 14.4 ± 6.3 years; age range = 5.9–31.8; 20.7% female; mean FIQ ± SD = 113.7 ± 13.5). Of these individuals, n=16 autistic and n=4 TD were excluded due to poor fMRI data quality (e.g., poor co-registration) following preprocessing (see *Neuroimaging data analysis* section). From the GENDAAR dataset, n=85 autistic individuals (mean age ± SD = 12.7 ± 2.9 years; age range = 8.0–18.0; 47.5% female; mean FIQ ± SD = 100 ± 20.3) were included based on the availability of both IQ and adaptive behavior measures as well as resting-state fMRI data and matched with n=76 TD controls (mean age ± SD = 13.3 ± 2.9 years; age range = 8.1–17.9; 48.0% female; mean FIQ ± SD = 112.1 ± 15.4). Following visual inspection after fMRI preprocessing, n=7 autistic and n=4 TD participants were excluded. An additional n=23 autistic and n=7 TD participants were excluded due to insufficient artifact attenuation by wavelet denoising as indicated by persistent DVARS spikes as judged by visual inspection. Lastly, 8 autistic and 11 TD participants were excluded due to excessive head motion (mean framewise displacement [FD] > 0.5 mm [39]). The final combined fMRI sample comprised n=174 autistic individuals (mean age ± SD = 12.3 ± 6.2 years; age range = 5.2–39.0; 24.1% female; mean FIQ ± SD = 106.9 ± 18.2) and n=185 TD controls (mean age ± SD = 14.3 ± 5.5 years; age range = 5.9–31.7; 30.2% female; mean FIQ ± SD = 113.1 ± 14.1).

Because ABIDE and GENDAAR are multisite imaging datasets, the MRI machines and scan parameters vary according to collection site. While all MRI machines used for data were at 3T field strength and were manufactured by Siemens, the actual model of the machine varied (e.g., Tim Trio, Prisma, Allegra). The repetition time (TR) and echo time (TE) for all sites was consistent at TR = 2000 ms and TE = 30 ms. Slice thickness varied from 3-4mm. The length of the functional scan sessions also varied between 165 to 180 volumes. All sites had data acquired under eyes open conditions, except for NYU2, which was eyes closed. For specifics of scan parameters for each collection site, see Supplementary Table 4. Because of this between site variability and known variance accounted for by collection site in multi-site imaging datasets, we utilize batch correction (i.e. ComBat) to account for these differences (see section on Neuroimaging data analysis for more details).

### Standardized clinical behavioral measures utilized for stratification

#### Vineland Scales of Adaptive Behavior (VABS)

The Vineland Adaptive Behavior Scales (VABS)*[26, 27]* is a widely used, standardized, semi-structured caregiver interview designed to assess adaptive functioning across the lifespan in both typical and clinical populations. For individuals aged 68 months and older, the VABS evaluates adaptive functioning across three domains: communication, daily living skills, and socialization. In this study, we used standardized scores for each domain, with a normative mean of 100 and a standard deviation of 15, indicating performance relative to age-appropriate typically developing norms. Three versions of the VABS were available in NDA: the VABS-II Parent Rating Form (n = 141), the VABS-II Interview Survey (n = 66), and the VABS-III (n = 212) (see Table 2 for additional details). In the resting-state neuroimaging dataset, n=127 participants had data from the VABS-II Parent Rating Form and n=47 from the VABS-II Interview Survey.

#### Wechsler Intelligence Scale for Children (WISC)

The Wechsler Intelligence Scale for Children (WISC)[28–30] is a widely used measure of cognitive ability for children aged 6 to 16 years (Wechsler et al., 2003). It comprises a series of subtests that yield both a full-scale IQ and indices reflecting specific cognitive domains. For the purposes of this work, we extracted an index of verbal intelligence (VIQ), corresponding to the Verbal Comprehension Index, and an index of non-verbal intelligence (NVIQ). The NVIQ is referred to as the Perceptual Organization Index in the WISC-III, the Perceptual Reasoning Index in the WISC-IV, and the average of the Fluid Reasoning and Visual Spatial Indices in the WISC-V. Within NDA, n=11 participants completed the WISC-IV and n=20 the WISC-V. No participant in the resting-state neuroimaging dataset had available WISC data.

#### Wechsler Adult Intelligence Scale (WAIS)

The Wechsler Adult Intelligence Scale (WAIS)[31, 32] is the standard instrument for assessing intelligence in individuals aged 16 years and older. Its structure mirrors that of the WISC but incorporates more complex items suitable for adult populations. For both the WAIS-III and WAIS-IV, we used the Verbal Comprehension Index as a measure of VIQ. The NVIQ was derived from the Perceptual Organization Index in the WAIS-III and from the Perceptual Reasoning Index in the WAIS-IV. Within NDA, n=26 individuals completed the WAIS-III and n=25 the WAIS-IV. No participant in the resting-state neuroimaging dataset had available WAIS data.

#### Wechsler Abbreviated Scale of Intelligence (WASI)

The Wechsler Abbreviated Scale of Intelligence (WASI)[33] is a brief and reliable measure of cognitive ability that provides a quick estimate of general intellectual functioning, VIQ and N-VIQ in individuals aged 6 to 90 years. Widely used in research settings, it includes only four subtests and serves as an efficient alternative to WISC and WAIS. For subjects whose IQ was measured with the WASI-II, we included both the Verbal Comprehension Index and the Perceptual Reasoning Index, respectively corresponding to VIQ and N-VIQ. Within NDA, n=188 autistic individuals completed the WASI-II, while in the resting-state neuroimaging dataset, n=92 participants had available WASI-II data.

#### Differential Abilities Scales (DAS-II)

The Differential Abilities Scale (DAS-II)[34] is a standardized intelligence test designed to assess cognitive functioning in children and adolescents. It includes two developmental ranges: DAS-II Early Years, for ages 2 years 6 months to 6 years 11 months, and DAS-II School-Age, for ages 7 years to 17 years 11 months. For this study, we extracted the Verbal Reasoning and Non-Verbal Reasoning indices, both standardized with a mean of 100 and a standard deviation of 15. Within NDA, n=188 individuals completed the DAS-II School-Age and n=31 the DAS-II Early Years. In the resting-state neuroimaging dataset, n=47 had DAS-II School-Age and n=27 DAS-II Early Years data.

### Batch correction for different clinical test versions

Before doing the primary downstream *reval* clustering analysis on NDA data, we had to address batch effects caused by different test versions of intelligence and adaptive functioning measures. For each of the VABS domains, we applied a batch correction factor to adjust the raw data. These factors correspond to the beta coefficients estimated in a linear model with VABS standard scores as the dependent variable and VABS version as the independent variable with age and sex as covariates. Batch correction factors are taken from Mandelli et al., [40] since they were estimated in a bigger sample size within the age range of interest for this work (*n* = 2,561; 6–61 year). To project out batch-related variability brought on by the VABS version, the correction factors (i.e. beta coefficients) were multiplied against the raw data and then the difference was removed from the raw VABS domain standard scores. To similarly correct for different intelligence test versions, we ran a linear model with verbal and nonverbal IQ scores from all the available cognitive tests data in NDAR (n = 1851) as dependent variables and test versions as independent variables with age and sex as covariates. Betas associated with each test version were then used to project out from IQ scores variance associated with IQ versions. Subsequent *reval* clustering analyses were carried out with version-corrected VABS and IQ data.

### Stability-based relative clustering validation analysis (reval)

Our stratification model utilizes an unsupervised data-driven clustering approach designed to identify the optimal number of clusters with the best level of generalizability in new data. This approach is based on stability-based relative clustering validation, implemented using our Python library *reval*[41, 42]. The core principle behind *reval* is to transform an unsupervised clustering framework into a classification problem, allowing direct evaluation of clustering reproducibility and generalizability. The *reval* algorithm solves for 2 goals – 1) identify the optimal number of clusters (k) and then 2) test a classifier trained on that optimal clustering solution to see if it can predict with high levels of accuracy the clustering labels identified in a held-out validation set. For more detail about *reval,* see Landi et al., (2021)[41]. Further examples of implementation of *reval* for similar kinds of stratification goals in autism can be found in Mandelli et al. (2023) [40], and Mandelli, Severino et al. (2024) [10]. For a schematic visual depiction of *reval*, see Figure 1.

**Figure 1.**
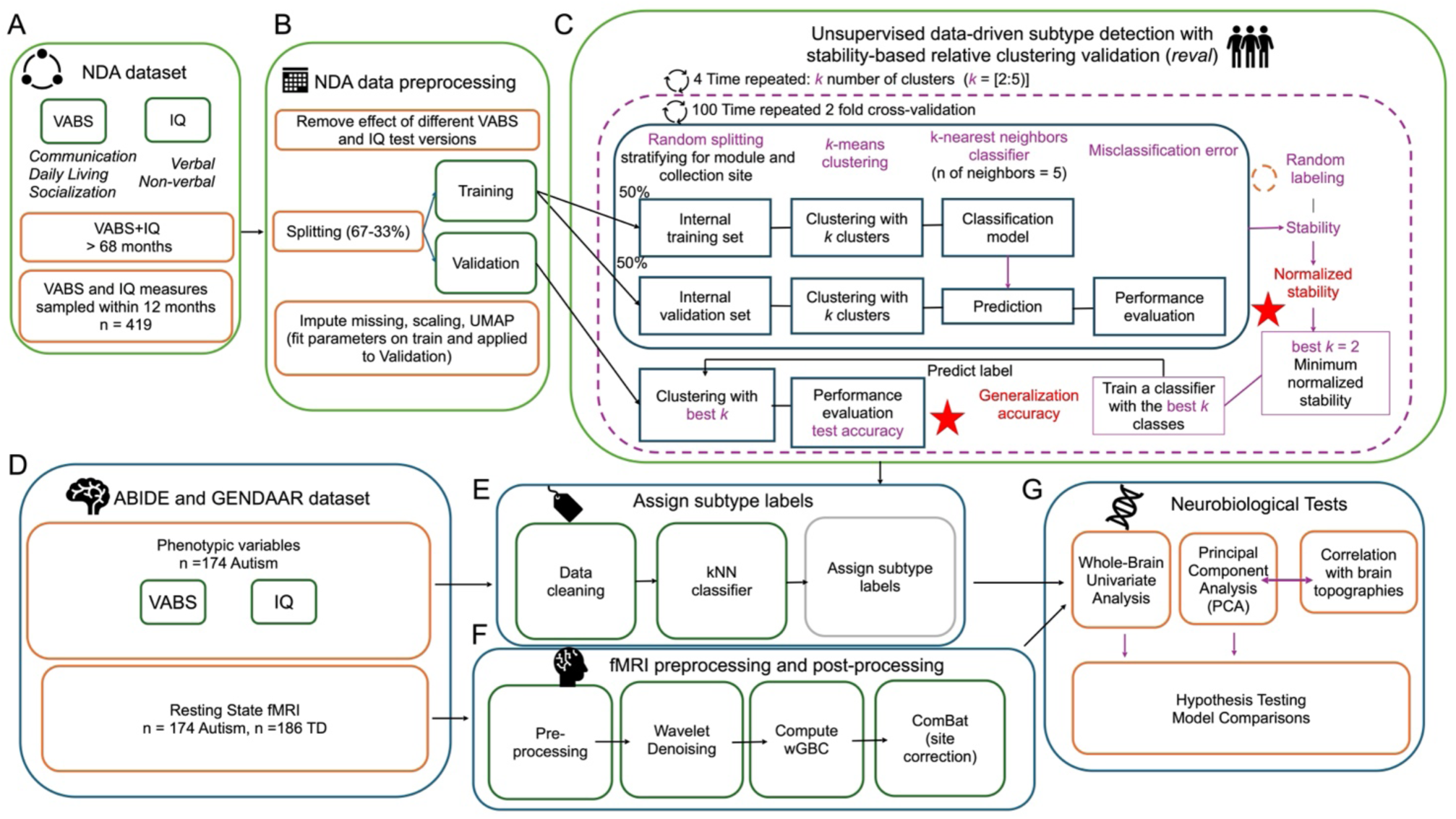
Data analysis workflow. This figure shows a workflow overview of the analyses conducted in this work. Panels A and B show how we select, clean, and preprocess data from the National Institute of Mental Health Data Archive (NDA). To identify subtypes in a data-driven manner, we used stability-based relative clustering validation analysis (reval) (panel C) on a final sample of n=419 autistic individuals. Panel D and E depict how we selected, cleaned, and labeled individuals by subtype from the neuroimaging datasets. Panel F show the main steps in the imaging analyses (i.e. preprocessing, denoising, wGBC computation, batch correction). Panel G illustrates the main univariate and multivariate analyses applied to test the neurobiological sensitivity of our stratification model.

Before entering the *reval* clustering analysis, NDA data was split into independent training and validation sets (67-33% split for training-validation) while balancing the split for variables such as age, sex, VABS version and intelligence test type. Two preprocessing steps are then applied – 1) scaling each feature in the dataset to mean of 0, standard deviation (SD) of 1 (*sklearn.preprocessing.StandardScaler*) and 2) dimensionality reduction from 5 to 2 features via Uniform Manifold Approximation and Projection (UMAP)[42] (https://umap-learn.readthedocs.io/en/latest; n_neighbors = 30, min_dist = 0.0, n_components = 2, random_state = 42, metric = Euclidean). Those preprocessing steps were implemented by fitting the parameters to the training set and then applying them to the validation set, thus preserving the independence between the training and validation sets. After preprocessing, the *reval* algorithm was run using k-means as the clustering algorithm and k-nearest neighbors (k-NN) as the classifier. To identify the optimal number of clusters (k), an internal 2-fold cross-validation scheme with 100 repeated iterations was applied to the training set. The goal of this internal cross-validation loop on the training set to identify optimal number of clusters. Clustering on each iteration is done through a range of clustering solutions between k=2 to k=5. Within the cross-validation loop on the training set, the algorithm clusters the internal train and test sets independently and then trains a classifier on the internal train set and then tests its accuracy at predicting the clustering labels determined in the internal test set. To account for the non-uniqueness of clustering labels, *reval* permutes the cluster labels and uses the Kuhn-Munkres algorithm to identify the labeling solution that minimizes misclassification error. This measure of misclassification error is called ‘clustering stability’[42] and ranges from 0–1, with lower values indicating more stable and reproducible clustering solutions. This measure of stability is then normalized to the stability of random labelings (100 random permutations) to arrive at the final measure of ‘normalized stability’. The clustering solution, k, with the lowest normalized stability within the internal cross-validation loop on the training set is then determined as the optimal k to use in further analysis. Once the optimal k has been identified, *reval* then trains a classifier on the entire training set to detect the optimal k solution. Clustering is then independently applied to the held-out validation set at the optimal k level, and then the classifier fit to the training set is applied to the validation set and the predicted versus actual clustering labels in the validation set are compared to arrive at a metric of generalization accuracy.

While the *reval* clustering allows for identification and estimation of how stable and reproducible a clustering solution is, it does not address the issue of whether actual true clusters exist. To test if actual true clusters exist, we perform a hypothesis test on the null hypothesis that no clusters are apparent in the data via a Monte Carlo simulation framework called SigClust. SigClust was designed to test whether clusters originate from a single multivariate Gaussian null distribution. SigClust is implemented with the *sigclust* [43] library in R with number of simulations set to 10,000. On each iteration of the simulation, SigClust generates a single multivariate Gaussian null distribution, applies k-means clustering at a specific optimal k, and then estimates a test statistic, CI, operationalized as the ratio of within-class sum of squares divided by the total sum of squares relative to the mean. After all iterations of the simulation are complete, the actual CI from the real data is compared to the CI simulated under the null hypothesis of a single multivariate Gaussian null. This allows for a p-value to be computed as the proportion of CI values from the simulated null distribution that are as small or smaller than the actual CI from the real dataset. To generate subtype labels in the imaging datasets we utilized a classifier fit to NDA data to identify the optimal k from *reval*. Here we utilized two types of subtyping models: 1) the VABS+IQ subtype model from the current work and 2) a VABS-only stratification model that comes from Mandelli et al., 2023 [40].

### Neuroimaging data analysis

Resting-state fMRI data across all sites followed the same preprocessing pipeline. This consisted of two components - core preprocessing and denoising. Core preprocessing was implemented with AFNI[44](http://afni.nimh.nih.gov/) using the tool speedypp.py [45] (http://bit.ly/23u2vZp).This core preprocessing pipeline included the following steps: (i) slice acquisition correction using heptic (7th order) Lagrange polynomial interpolation; (ii) rigid-body head movement correction to the first frame of data, using quintic (5th order) polynomial interpolation to estimate the realignment parameters (3 displacements and three rotations); (iii) obliquity transform to the structural image; (iv) affine co-registration to the skull-stripped structural image using a gray matter mask; (v) nonlinear warping to MNI space (MNI152 template) with AFNI 3dQwarp; (vi) spatial smoothing (6 mm FWHM); and (vii) a within-run intensity normalization to a whole-brain median of 1000. Core preprocessing was followed by denoising steps to further remove motion-related and other artifacts. Denoising steps included: (viii) wavelet time series despiking (‘wavelet denoising’); (ix) confound signal regression including the six motion parameters estimated in (ii), their first order temporal derivatives, and ventricular cerebrospinal fluid (CSF) signal (referred to as 13-parameter regression). The wavelet denoising method has been shown to mitigate substantial spatial and temporal heterogeneity in motion-related artifact that manifests linearly or non-linearly and can do so without the need for data scrubbing[46]. Wavelet denoising is implemented with the Brain Wavelet toolbox (http://www.brainwavelet.org). The 13-parameter regression of motion and CSF signals was achieved using AFNI 3dBandpass with the –ort argument. To further characterize motion and its impact on the data, we computed mean frame displacement (FD) and standard deviation of successive difference image (DVARS)[39]. Individuals were excluded if mean FD exclusion was greater than 0.5 mm. Two of the authors (IS and MVL) also quality checked all data to ensure that the data included comprised individuals whose motion-related DVARS spikes before denoising were heavily attenuated after wavelet denoising. After these data exclusions and manual QC procedures we still found that mean FD could be explained by significant main effects of subtype (VABS+IQ: F(2,356) = 10.40, *p* = 4.12 × 10⁻⁵; VABS-only: F(2,356) = 11.63, *p* = 1.30 × 10⁻⁵) and acquisition site (F (6, 352) = 4.17, *p* = 4.67 × 10⁻⁴) (Supplementary Figure 1). Therefore, further procedures implemented after wGBC was computed, were used to mitigate these issues (e.g., projecting out principal components heavily saturated by head motion effects, batch correction, covariate adjustment).

After preprocessing, we parcellated the data into 180 regions defined by the symmetric Human Connectome Project Multimodal Parcellation (HCP-MMP)[47]. Weighted global brain connectivity (wGBC) was then computed on each parcel and can be defined as the average correlation between a seed parcel and all other brain parcels[48]. In graph theory, wGBC is equivalent to weighted degree centrality. After estimating wGBC, we investigated whether site-related batch would account for variance in the data. Site-related effects on wGBC were corrected for using the batch correction technique called ComBat [49], implemented within the ’sva’ package[50] in R (https://www.bioconductor.org/packages/release/bioc/html/sva.html). The model for ComBat had wGBC as the dependent variable and had independent variables of site (as the batch effect), mean FD, sex, age, subtype, and the interaction between sex*subtype and age*subtype. To assess the effectiveness of ComBat, we performed principal components analysis (PCA) before and after ComBat adjustment. As expected, we found that ComBat reduced the prominent site effect observed on the first PC (Supplementary Figure 2). However, after batch correction, we found that the first PC was still highly sensitive to the effect of head motion (mean FD). Therefore, we projected out variance from this first head-motion explained PC as a final denoising step to ensure that head motion-related effects on wGBC are mitigated. After all these analysis steps, we utilized k-nearest neighbors (kNN) imputation, where the k parameter is set to 3 neighbors, to handle a small number of individuals with some missing brain regions due to variable/poor coverage. This kNN imputation procedure was implemented on n=17 brain regions and was done only in situations to patch up missing values when the region with a missing value met a criterion of having <10% of individuals from the sample having missing values. Regions with >10% of individuals with missing data (n = 1 region; TGv in the anterior temporal lobe) were excluded from further analysis.

### Mass univariate analysis versus multivariate cortical patterning analysis on wGBC

We took two approaches to assessing group differences in wGBC – 1) a mass univariate whole-brain approach and 2) a multivariate cortical patterning approach. For the mass univariate approach, we simply ran hypothesis tests separately on every parcel in the HCP-MMP. The models for these hypothesis tests were general linear models (GLM) with a region’s wGBC as the dependent variable and mean FD, eye condition (i.e. open or closed), sex, age, subtype, and the interaction of subtype*age and subtype*sex as the independent variables. Effects were considered significant if they passed a multiple comparison correction over all 180 parcels of a false discovery rate (FDR) of q<0.05.

In contrast to mass univariate hypothesis testing, we also implemented analyses that were positioned to allow us to assess multivariate cortical patterning. These analyses utilized principal components analysis (PCA) to isolate latent variables that summarize how wGBC is varies across the brain in a patterned fashion. The input to PCA was a wGBC matrix with subjects along the rows and HCP-MMP parcels along the columns. Given our specific hypotheses for testing the S-A axis, we then identified components from the PCA that statistically matched the S-A axis via correlation between each PC’s loading coefficient maps and the S-A ranking map taken directly from Sydnor et al., (2021)[51] (https://github.com/PennLINC/S-A_ArchetypalAxis). We also tested if PC maps would be similar to genomically sensitive maps, via computing the correlation between wGBC PC maps and the first principal component of adult gene expression data from the Allen Human Brain Atlas (AHBA PC1) [52,53]. The *neuromaps* Python library [53] (https://github.com/netneurolab/neuromaps?tab=readme-ov-file) was used to extract AHBA PC1 parcellated by the HCP-MMP. Statistical comparisons between the wGBC maps and the S-A axis or AHBA maps were implemented by computing Spearman ρ and then evaluating statistical significance via permutation p-values from ‘spin tests’ (10,000 repetitions) that preserve spatial autocorrelation [54–56]. After identifying PCs that matched the S-A axis, we then performed similar GLM models using the PC scores as the dependent variable and using mean FD, eye condition (i.e. open or closed), sex, age, subtype, and the interaction of subtype*age and subtype*sex as the independent variables.

### Data and code availability

Tidy data and code for this work is publicly available at https://gitlab.iit.it/bmp006-public/LAND_IIT/a3d_vabsiq_wgbc.

## Results

### Detection of autism phenotypic subtypes

The first objective of this work was to test whether it is possible to detect autism subtypes based on non-core LIAF features. Stability-based relative clustering validation (*reval*) [41] on IQ and VABS data from NDA identified a two-cluster solution as the optimal clustering solution. This solution can be turned into a classifier and then utilized to predict clustering labels in an independent validation set. We find that the classifier can predict clustering labels in an independent validation set with very high generalization accuracy (i.e. 92%) (Fig. 2a). SigClust analysis confirms that this solution is indeed indicative of true clusters that heavily deviate from a single multivariate Gaussian null distribution (*p* = 9.99e-4). The two clusters are shown in Fig 2b-c and can be described as a distinction of individuals that are relatively ‘*High*’ versus ‘*Low*’ across all features. Notably, there is no hard cutoff between the subtypes. While the distributions of the subtypes overlap to some extent, effect sizes remain rather large throughout (*Cohen’s d* > 1.26) (Supplementary Table 3).

**Figure 2.**
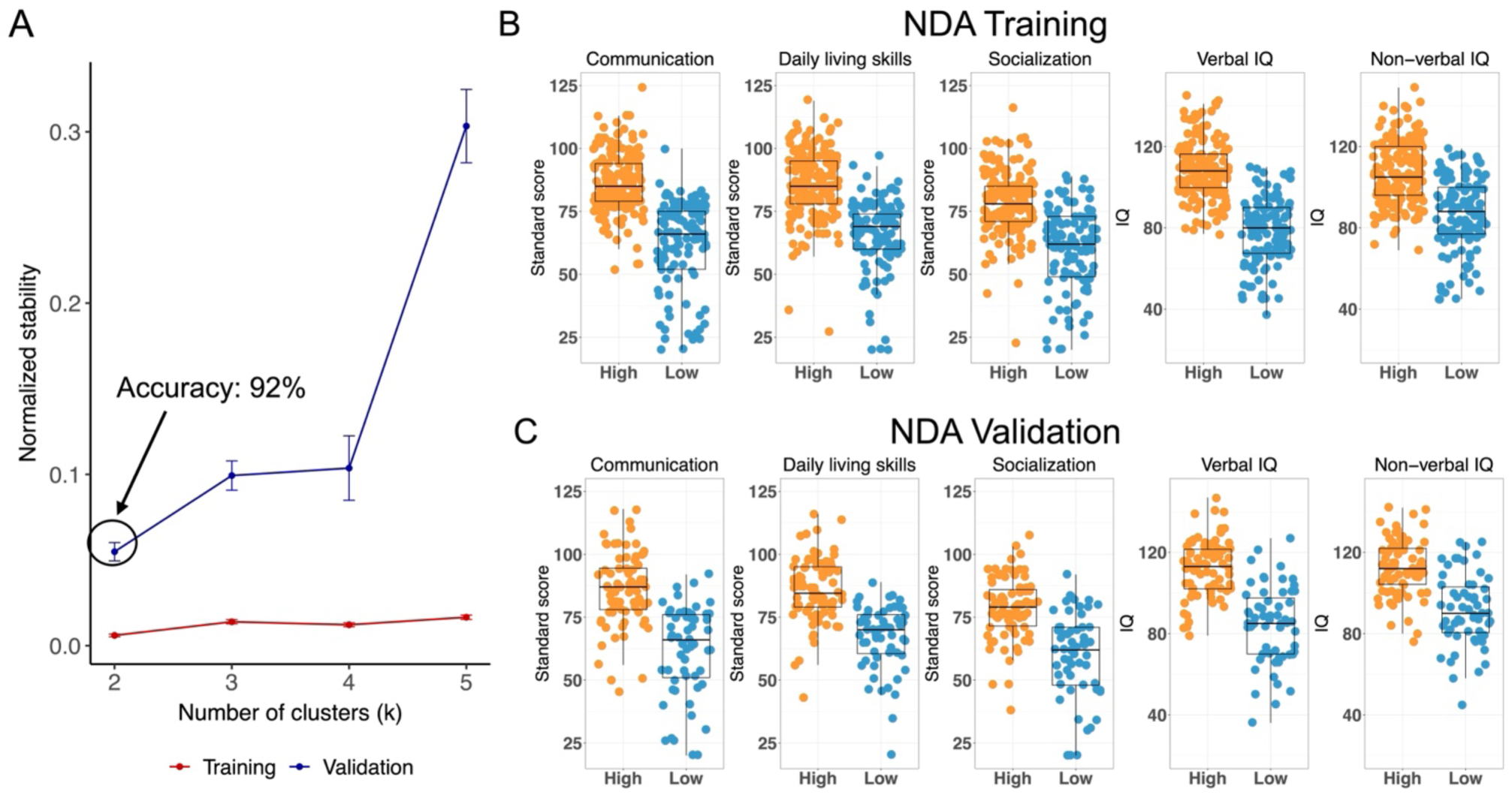
Identification of autism subtypes. Stability-based clustering validation (reval) detects two autism subtypes characterized by different levels of language, intellectual, and adaptive functioning (LIAF) features. Panel A shows that a 2-subtype solution is the optimal clustering solution that minimizes normalized stability and generalizes with 92% accuracy in the independent NDA validation dataset. Panels B-C plots IQ and VABS scores for the NDA Training (B) and NDA Validation (C) datasets. The two subtypes manifest as relatively high (High; orange) versus low (Low; blue) scores across all features.

### Autism stratification model comparison

Given the high levels of generalization accuracy from the NDA subtype model, we generated a subtype prediction model from NDA and then applied it to IQ and VABS data from independent resting state neuroimaging datasets. Here we examined two different kinds of subtype models – 1) the VABS+IQ model reported above, and 2) a VABS-only model derived from NDA and which has been previously published [40]. Examining these two models allows for subtype model comparisons when subtypes are defined by VABS-only versus when the subtype model includes information from both VABS and IQ measures (i.e. VABS+IQ). Of the n=174 autistic individuals in the neuroimaging dataset, n=118 were classified as *High* and n=56 as *Low* based on the VABS+IQ subtype model (Fig. 3a). In contrast, the VABS-only subtype model classified n=141 individuals as *High* and n=33 as *Low*. Statistical comparison of these two models indicates that the VABS-only subtype model was significantly more conservative in assigning individuals to the *Low* subtype compared to the VABS+IQ subtype model (*χ²* = 30.67, *p* = 3.06e-8) (Fig. 3a). Next we examined whether autism subtypes differed in terms of their demographic characteristics. Descriptive statistics for age and sex are reported in Supplementary Table 5. In both VABS+IQ and VABS-only subtype models, there were no significant differences in the proportion of males to females between subtypes, with a male:female ratio of approximately 2.9:1 (VABS+IQ : *χ²* = 0.89, *p* = 0.35; VABS model: *χ²* = 1.88, *p* = 0.17) which aligns relatively well with the ratio reported in the general autism population[57]. In contrast, subtypes significantly differed in age at scan in both subtype models (VABS+IQ: *F(2,356)* = 7.00, *p* = 0.001; VABS-only *F(2,356)* = 11.17, *p* = 9.65e-5). Within the VABS+IQ model, the *High* subtype was significantly younger than the TD group (*t(356)* = -3.73, *p* = 0.0007). Within the VABS-only model, the *High* subtype was younger than the TD group (*t(356)* = -4.18, *p* = 0.0001) and the *Low* subtype (*t(356)* = -3.46, *p* = 0.002) (Supplementary Table 5). To account for these differences between subtypes in age at scan, we included age at scan as a covariate in all subsequent analyses.

**Figure 3.**
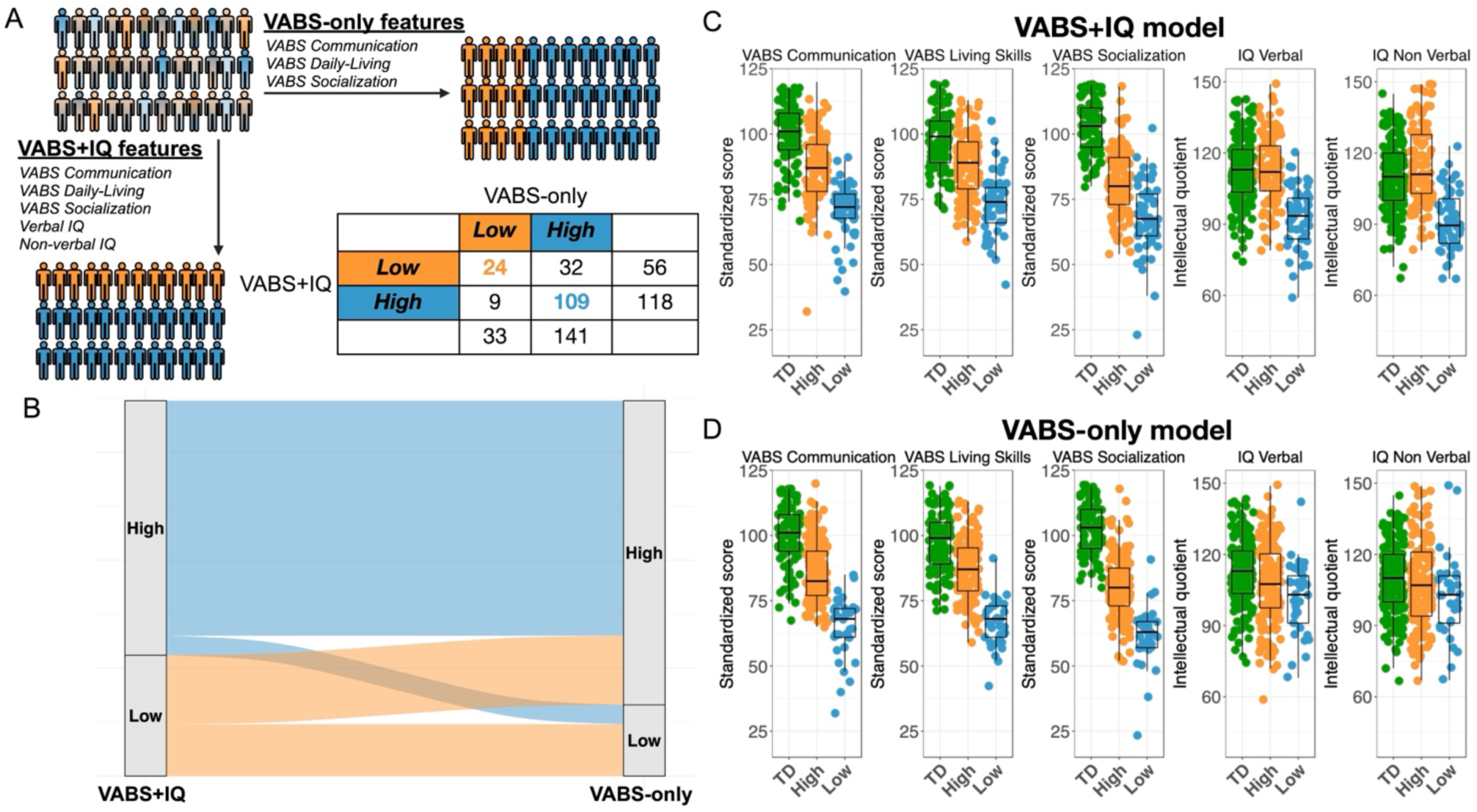
Comparison between VABS+IQ and VABS-only subtype models. Autistic individuals from the ABIDE and GENDAAR datasets were classified into distinct autism subtypes using two models: the VABS+IQ model reported in this work and a second model based solely on Vineland domains (i.e. VABS-only)[40]. Panel A displays a breakdown of the sample sizes in the imaging dataset for each subtype within each subtype model. Panel B presents an alluvial plot illustrating how individuals stay or shift between Low and High subtypes in each of the two models. Panels C and D show scatter-boxplots of data across all features and groups.

### Mass univariate analysis of subtype differences in global brain connectivity

In our first analysis we utilized a mass univariate approach to search throughout the entire brain and identify any possible differences between subtypes in wGBC. This analysis resulted in very few significant differences. In both stratification models, only two brain regions survive showing a significant difference between subtypes after FDR correction. These regions are a visual association area (V6 VABS+IQ: *F(2,348*) = 8.30, *p* = 0.0004; VABS-only: *F(2,348*) = 11.34, *p* = 1.56e-5) and an area within the anterior cingulate cortex (d23ab VABS+IQ: *F(2,348*) = 7.88, *p* = 0.0003; VABS-only: *F(2,348*) = 8.29, *p* = 0.0003). In both stratification models, *High* and *Low* autism subtypes showed consistent patterns of hypo-connectivity in V6 and hyper-connectivity in d23ab relative to the TD group (Supplementary Figure 3). This shared connectivity profile across subtypes suggests a prominent case-control effect exists, although effect sizes were more pronounced for the TD vs. *Low* comparison than for TD vs. *High* Supplementary Figure 3). Results of mass univariate analyses are summarized in Supplementary Table 6-7.

### Subtype differences in patterning of connectivity along the sensorimotor-association axis

Next, we examined whether the distinction between VABS+IQ and VABS-only subtypes could be better captured as a difference in multivariate patterning of connectivity across the cortex. We hypothesized that a specific type of wGBC patterning varying along the important S-A axis might be most sensitive at differentiating the subtypes. Using PCA, we first decomposed wGBC into orthogonal PCs that reflect how connectivity is patterned across the cortex. We then compared each wGBC PC coefficient map to the S-A axis to identify which components, if any, may match the S-A axis. This analysis identified wGBC PC1 as the sole PC with a strong association with the S-A axis (VABS+IQ: *ρ* = 0.78, *p_SPIN_* = 9.99e-4; VABS-only: *ρ* = 0.77, *p_SPIN_* = 9.99e-4) (Figure 4D). Additional analysis also revealed that wGBC PC1 was also highly similar to the main axis of gene expression variation in the adult human brain (AHBA PC1 VABS+IQ: *ρ* = 0.55, *p_SPIN_* = 0.038); VABS-only: *ρ* = 0.55, *p_SPIN_* = 0.048) (Figure 4E).

**Figure 4.**
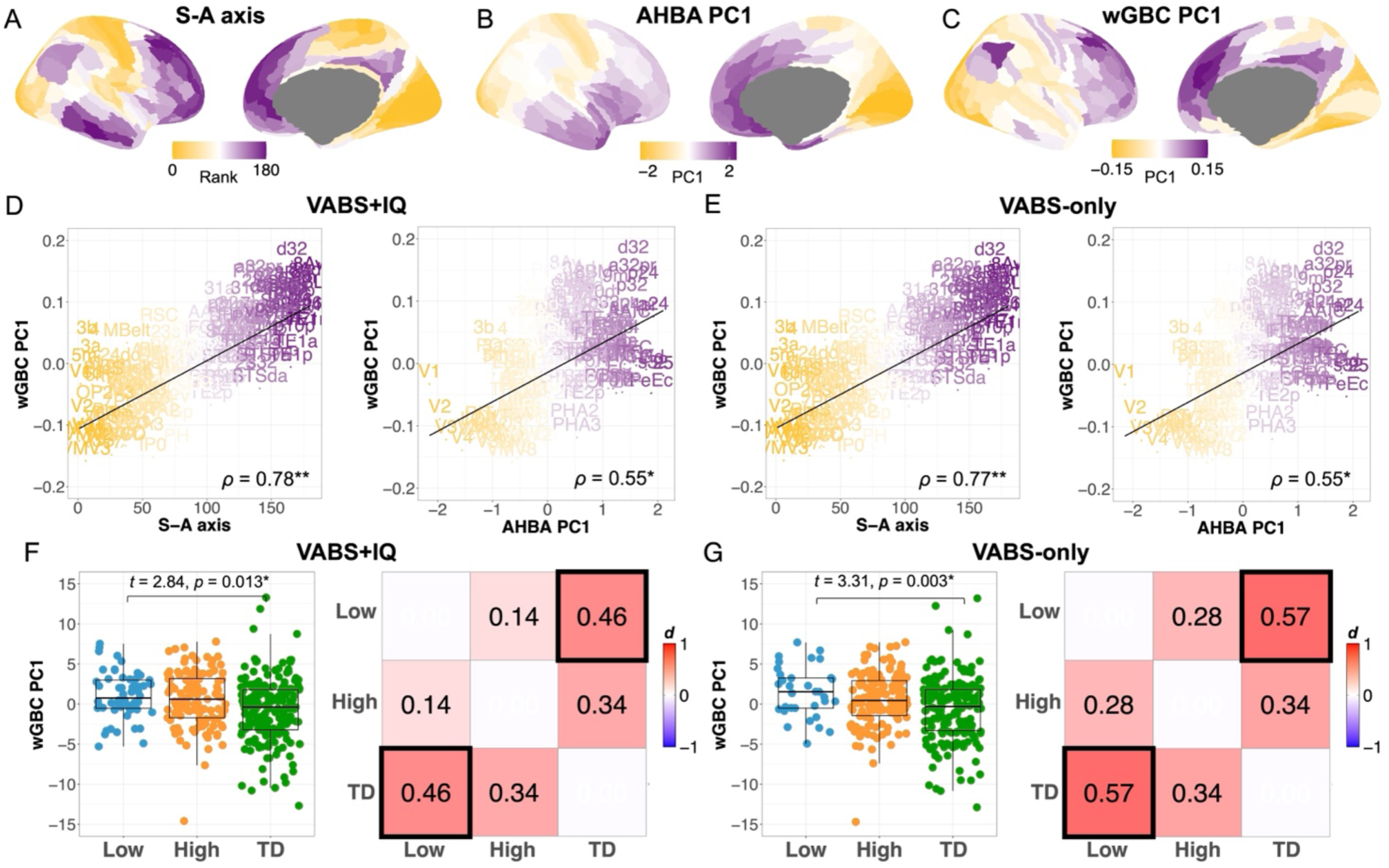
Multivariate cortical patterning of wGBC. Panel a shows cortical patterning along the sensorimotor–association axis (S-A axis; Sydnor et al., 2021). Panel b shows the first principal of gene expression in the human adult brain from the Allen Human Brain Atlas (AHBA PC1; Hawrylycz et al., 2012). Panel c shows cortical patterning of wGBC derived from the current work. Panels d-e show scatterplots of S-A rankings (x-axis) or AHBA PC1 weights (x-axis) against wGBC PC1 (y-axis) for the VABS+IQ (C) and VABS-only (D) models. Panels f-g shows scatter-boxplots (left) and standardized effect sizes (Cohen’s d) heatmaps (right) for the VABS+IQ (E) and VABS-only (F) subtype models. The main effect of subtype on PC1 scores is primarily driven by differences between the Low subtype and the typically developing (TD) group. One asterisk (*) indicates a p_SPIN_<0.05, while two asterisks (**) indicates a p_SPIN_ < 0.0001.

Having isolated PC1 as the main wGBC patterning effect varying along the S-A axis, we next followed up these analyses with specific hypothesis tests on wGBC PC1. Here we find a main effect of subtype for both VABS+IQ (*F(2,348)* = 4.66, *p* = 0.010) and VABS-only models (*F(2,348)* = 5.56, *p* = 0.004). With further pairwise comparisons between each group we find that these subtype effects are mainly driven by the pairwise comparison between *Low* and TD, with the *Low* subtype having significantly higher PC scores compared to the TD group (VABS+IQ: *t(348)* = 2.84, *p* = 0.013, *Cohen’s d* = 0.46; VABS-only: *t(348)* = 3.31, *p* = 0.003, *Cohen’s d* = 0.57) (Fig. 4F-G). Since PC scores themselves may not adequately reflect the true directionality of the group differences, we reconstructed the data for PC1 back to the original space in order to interpret the directionality of these group differences. Here we find that the patterning difference in connectivity is due to hyper-connectivity in the *Low* subtype compared to TD for association areas, but hypo-connectivity in *Low* compared to TD for sensorimotor areas. Finally, we directly compared VABS+IQ and VABS-only subtype models using the Akaike Information Criterion (AIC). This analysis would allow us to see whether one of the subtype models is sufficiently better than the other at explaining variance in wGBC effects. Here we find that AIC values were largely similar for both models (VABS+IQ: *AIC* = 1949.95; VABS-only: *AIC* = 1945.95; *ΔAIC* = 3.99) which suggests that both models are likely equally sensitive at detecting differences between subtypes.

## Discussion

In this study we find that an autism subtype characterized by lower levels of language, intellectual functioning, and adaptive functioning (LIAF) exhibits altered global functional connectivity along the sensorimotor–association (S-A) axis [51]. Using a robust unsupervised subtyping approach, we identified two reproducible and generalizable subtypes within the autistic population from school-age to adulthood. These subtypes, defined by relatively lower versus higher abilities across LIAF domains, roughly correspond to previously proposed early developmental distinctions based on language, intellectual, motor, and adaptive functioning (LIMA) features [10]. Our findings demonstrate that the LIAF-based distinction captures a specific effect of cortical patterning of functional connectivity described by global hyper-connectivity of associative regions and global hypo-connectivity of sensorimotor areas in the lower-ability subtype. These cortical patterning effects on global functional connectivity that may align well with hierarchical brain organization along the well-known S-A axis as well as the underlying biological programs that regulate such patterning (e.g., gene expression variation) [16].

The current work builds on long-standing work examining the idea of atypical neural connectivity in autism [58], as well as the idea of describing autism as a set of ‘disconnection syndromes’ [59]. Several multidisciplinary and convergent approaches (e.g., genetic, post-mortem, animal model, neuroimaging) have validated that autism can be characterized by atypical neural connectivity (e.g., [24, 60–64]). Focusing on in-vivo neuroimaging studies, case-control studies have observed a mosaic pattern of both hyper- and hypo-connectivity in the autistic brain (e.g., [20, 21, 35, 65–67]) and some studies give hints to the idea that such mosaic patterning may align with the S-A axis (e.g., [20, 21]). This work extends such observations by potentially showing that such case-control effects may be driven by phenotypic variation aligning with LIAF features.

The significance of our findings that global functional connectivity differs in a LIAF autism subtype along the S-A axis of hierarchical brain organization is also important to discuss. The S-A axis is a major cortical axis for which multiple neurodevelopmentally sensitive phenomena unfold along. Most proximal to the data modality of interest here (rsfMRI), the S-A axis aligns closely with the functional gradients first observed by Margulies and colleagues [68]. However, the S-A axis is also sensitive to cortical gradient effects that stem outside the modality of rsfMRI. For example, in terms of structural cortical maturational effects, developmental cortical expansion, thickness, and allometric scaling [69–71] effects unfold along a similar axis. Anatomical hierarchical organization, as shown with the myelin-sensitive T1/T2 ratio, also varies along a similar axis [72]. The convergence of such structural and functional neurodevelopmentally sensitive effects may be latent phenomena emerging from early developmental events that set up and differentiate cortical regions to have their own molecular identities [11]. Cortical regions start to develop their unique molecular identities via transcriptomic programs that allow for a diverse range of cell types and circuits to form. Genomically there is evidence that the principal axis of gene expression variation in the adult brain follows the S-A axis [51, 52]. However, early in prenatal development, such cortical transcriptomic differentiation begins along anterior-posterior (A-P) and dorsal-ventral (D-V) gene expression gradients[73]. As individuals develop postnatally, activity-dependent gene expression programs further sculpt hierarchical patterning away from this A-P and D-V prenatal organization and into the more mature form of S-A organization present in the adult brain [16–18]. An atypical neurodevelopmental trajectory from A-P and D-V prenatal patterning to more mature S-A patterning may be a convergent circuit-level explanation for autism and some aspects of phenotypic heterogeneity within autism. Corroborating this idea is work showing that S-A axes are less functionally differentiated in autistic adults [19]. Thus, identification of the S-A gradient as a principal source of subtype differences in autism global functional connectivity may be telling of underlying neurobiology linked to how cortical regions hierarchically mature and develop their own identities.

The S-A axis has also been theorized to be an important for explaining differences in information processing in autism. Bernhardt and colleagues have recently suggested that the sensory-transmodal hierarchy of brain organization, along which the S-A axis tracks well, may help to explain a variety of heterogeneous cognitive and behavioral phenotypes in autism [22]. In particular, autism is notable for atypical cognitive phenotypes that span the gamut of sensory (e.g., sensory or perceptual processing) to transmodal (e.g., social cognition) brain circuitry. Bernhardt and colleagues suggest that the array of alterations in information processing may be explained by alterations in the underlying neural circuitry that spans the hierarchical sensory-transmodal axis of brain organization. The current work is compatible with this idea and adds a component by showing that the global functional connectivity profile over the entire brain can differ along the S-A axis and may co-occur with different LIAF phenotypes.

The current work also builds on an emerging literature that suggests that cortical patterning may be a key underlying neural mechanism that differentiates autisms with differing language, intellectual, motor, and adaptive functioning features (i.e. LIMA features). At the level of gene expression, several post-mortem studies have shown that brain regions are transcriptomically less distinctive in autism (e.g., [12–14]). Such patterning effects with gene expression also emerge when examining gene expression relationships with neuroimaging phenotypes (e.g., task-related fMRI response to speech, cortical thickness, surface area) [6, 7, 10]. Importantly, these studies on imaging-gene expression relationships highlight that the genomic cortical patterning effects on neuroimaging phenotypes are distinctive in individuals with the most affected LIMA/LIAF features. Indeed, these studies presented the basis for hypothesizing in the current study that cortical patterning of another neuroimaging phenotype (e.g., resting state global functional connectivity) would be atypical, particularly in a subtype with the most affected LIAF features. Thus, this work adds to a growing literature supporting a cortical patterning hypothesis that may underpin an autism subtype with poor early LIMA features and subsequently slower rates of developmental growth and potentially poorer later life outcomes.

We also compared classification approaches using models based on adaptive functioning alone (VABS) versus combined adaptive and intellectual functioning (VABS+IQ) and found that both approaches reveal similar patterns of neural differences. While the two approaches yielded modest differences in classification, the VABS model was more conservative in identifying individuals in the *Low* subtype. However, it is notable that most individuals examined with rsfMRI data and which were classified as *Low* in the VABS+IQ model had verbal and non-verbal IQ scores above the threshold for what is typically considered as prominent intellectual disability. This highlights a persistent challenge in autism neuroimaging research: the under-representation of lower intellectual functioning and minimally or non-verbal individuals, largely due to practical constraints in MRI data acquisition [74, 75]. Even in the situation that more MRI scanning data was available individuals with very low non-verbal IQ, it is notable that standard IQ assessments may fail to fully capture the complexity of intellectual functioning in some autistic individuals [76]. We suggest that addressing this inclusion bias in MRI studies will be critical to making the Low subtype more representative of the broader autistic population as this may help for potentially revealing even stronger neural differences associated with LIAF features.

There are several limitations and caveats to discuss in this work. First, in keeping with many studies showing effects of head motion on functional connectivity [21, 39, 77], a substantial amount of variance in preprocessed global functional connectivity measures were explained by head motion. We attempted to address this with a strict head motion threshold for inclusion, alongside employing denoising strategies such as wavelet and PCA denoising. Nevertheless, results should be replicated in future work with much lower head motion data and more advanced data collection and denoising strategies (e.g., [78]). Second, while our phenotypic stratification models were built without specific biases inherent to imaging studies, the application of our classification to MRI-specific ABIDE I, ABIDE II, and GENDAAR datasets may not be representative of autistic individuals with lower IQ and language abilities. Third, independent datasets for use in replication were not available in this work. Thus, future work in much larger samples should test for replication of the observed effects.

In summary, our findings demonstrate that autism subtypes defined by language, intellectual, and adaptive functioning (LIAF) correspond to distinct large-scale functional connectivity patterns along the sensorimotor–association axis. These results underscore the importance of the S-A axis in some phenotypic subsets of autistic individuals. Furthermore, the results suggest that underlying cortical patterning mechanisms may be altered in autistic individuals with low LIAF features. At a broader level, these results highlight the importance of phenotypic stratification of autism based on non-core phenotypic features and may offer a novel window into uncovering the neurobiological mechanisms that contribute to heterogeneity in autism.

## Data Availability

Data for this study comes from publicly available data from the Autism Brain Imaging Data Exchange (ABIDE; https://fcon_1000.projects.nitrc.org/indi/abide/) or from the National Institute of Mental Health Data Archive (NDA; https://nda.nih.gov). Tidy data and code for this work is publicly available at https://gitlab.iit.it/bmp006-public/LAND_IIT/a3d_vabsiq_wgbc.

https://nda.nih.gov

## Acknowledgments

We thank all participants and their families for participating in this study. We also thank Alessandro Gozzi and Jorge Jovicich for helpful discussions.

## Author Contributions

Conceptualization: MVL, IS, VM. Methodology: MVL, IS, VM. Formal analysis: MVL, IS, VM, NB. Investigation: MVL, IS, VM. Writing - original draft preparation: MVL, IS, VM. Writing - review and editing: MVL, IS, VM, NB. Visualization: MVL, IS, VM. Supervision: MVL. Project administration: MVL. Funding acquisition: MVL.

---

**Supplementary Figure 1.**
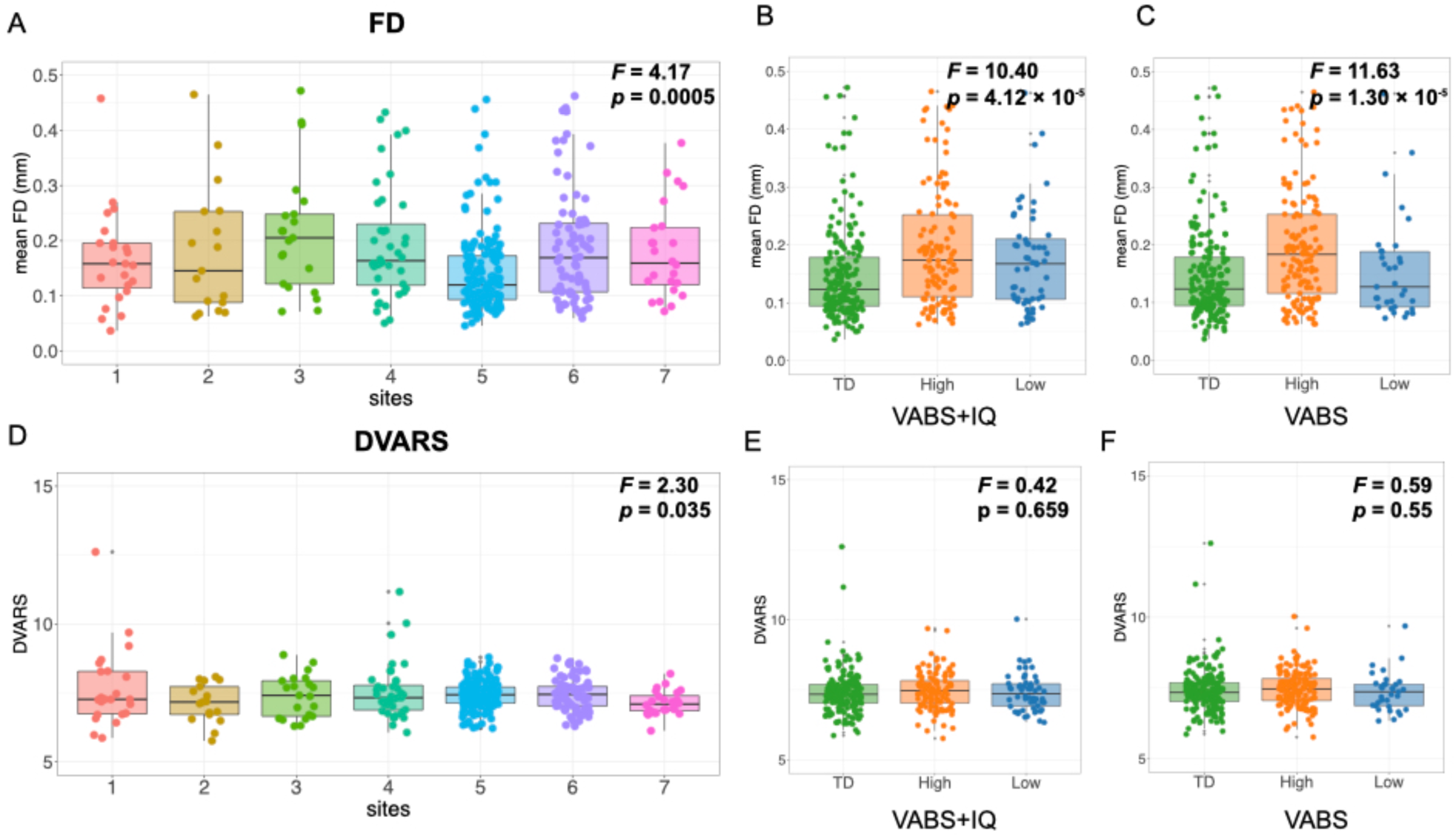
Mean framewise displacement (mean FD) and DVARS. Panel A shows differences in mean FD across sites. Panels B and C display mean FD differences between subtypes identified by the VABS+IQ and VABS-only models, respectively. Panel D shows DVARS across sites, while panels E and F report DVARS differences between subtypes for the two models.

**Supplementary Figure 2.**
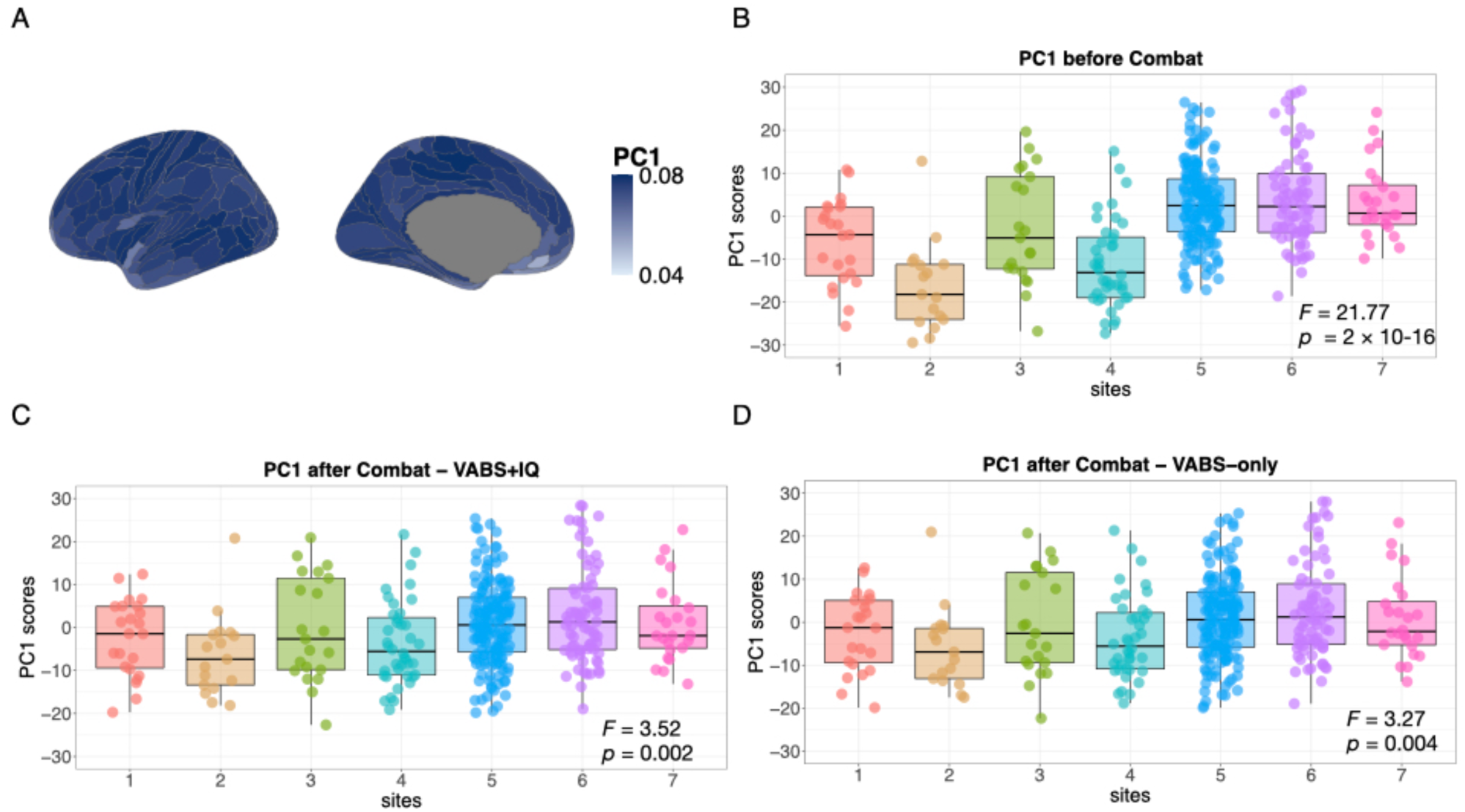
Effect of site-effect batch correction on wGBC. Panel A shows the loadings over all brain regions for the first principal component from a PCA on wGBC, before any site-related batch correction or other denoising has been implemented. This component explains ∼79% of the variance in wGBC. The plot shows a global effect of PC1 that is relatively similar across brain regions. Panel B shows PC1 scores over the 7 different sites before any site-effect batch correction was implemented. Considerable variation in PC1 scores exist between sites. Panels C–D show the PC1 scores after site-related batch correction was implemented for data from the VABS+IQ subtype model (C) or the VABS-only subtype model (D). Note the substantial reduction in variation between sites after batch correction compared to before batch correct (e.g., panel B).

**Supplementary Figure 3.**
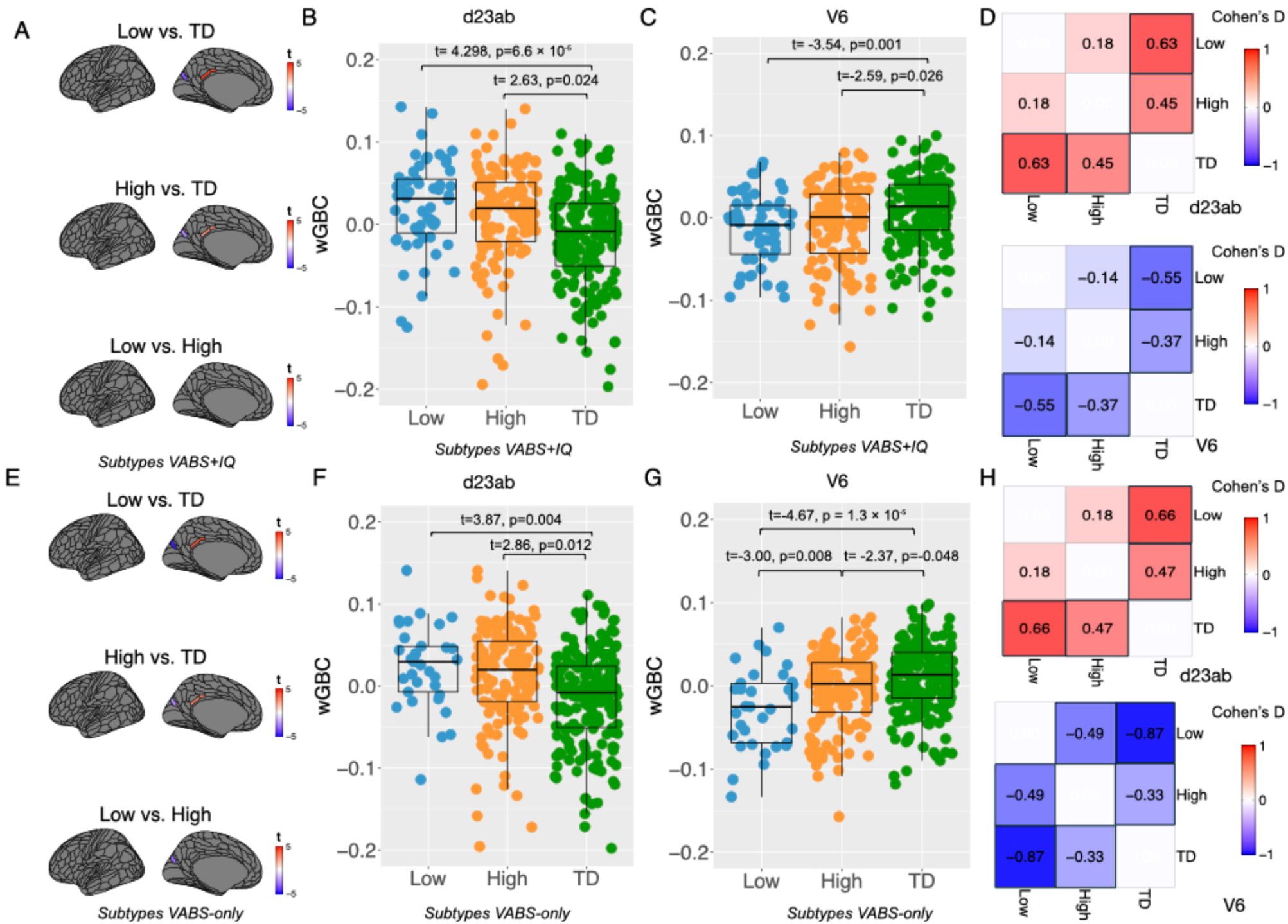
Summary of results from mass univariate analysis of wGBC. Two regions showed significant subtype effects after FDR correction (q < 0.05) in VABS+IQ (panels A-D) and VABS-only (panels E-H) subtype models. Region d23ab is located within the posterior cingulate cortex and, while region V6 is located within the occipital lobe. These regions are shown in panels A and E, with color representing t-statistics. Blue represents hypoconnectivity in autism subgroups relative to TD, while red indicates hyperconnectivity relative to the TD group. Panels B-C and F-G represent scatter-boxplots to show wGBC (y-axis) over groups (x-axis) for regions d23ab (left) and V6 (right). Panels D and H show effect size heatmaps for each region and all pairwise group comparisons.

